# Self-harm and the COVID-19 pandemic: a study of factors contributing to self-harm during lockdown restrictions

**DOI:** 10.1101/2020.12.04.20244129

**Authors:** Keith Hawton, Karen Lascelles, Fiona Brand, Deborah Casey, Liz Bale, Jennifer Ness, Samantha Kelly, Keith Waters

## Abstract

**Introduction:** The COVID-19 pandemic and resulting public health measures may have major impacts on mental health, including on self-harm. We have investigated what factors related to the pandemic influenced hospital presentations following self-harm during lockdown in England.

**Method:** Mental health clinicians assessing individuals aged 18 years and over presenting to hospitals in Oxford and Derby following self-harm during the period March 23^rd^ to 17^th^ May 2020 recorded whether the self-harm was related to the impact of COVID- 19 and, if so, what specific factors were relevant. These factors were organized into a classification scheme. Information was also collected on patients’ demographic characteristics, method of self-harm and suicide intent.

**Results:** Of 228 patients assessed, in 46.9% (N=107) COVID-19 and lockdown restrictions were identified as influencing self-harm. This applied more to females than males (53.5%, N=68/127 v 38.6%, N=39/101, χ^2^ = 5.03, p=0.025), but there were no differences in age, methods of self-harm or suicide intent between the two groups. The most frequent COVID-related factors were mental health issues, including new and worsening disorders, and cessation or reduction of services (including absence of face-to-face support), isolation and loneliness, reduced contact with key individuals, disruption to normal routine, and entrapment. Multiple, often inter- connected COVID-related factors were identified in many patients.

**Conclusions:** COVID-related factors were identified as influences in nearly half of individuals presenting to hospitals following self-harm in the period following introduction of lockdown restrictions. Females were particularly affected. The fact that mental health problems, including issues with delivery of care, predominated has implications for organisation of services during such periods. The contribution of isolation, loneliness and sense of entrapment highlight the need for relatives, friends and neighbours to be encouraged to reach out to others, especially those living alone. The classification of COVID-related factors can be used as an aide-memoire for clinicians.

## INTRODUCTION

The potential implications of the COVID-19 pandemic for mental health have been well articulated (Holmes et al, 2020), including the possible impacts on suicide and self-harm (Reger et al, 2020; Gunnell et al, 2020; Niederkrotenhaler et al, 2020). While increases in suicide and self-harm are not inevitable (Gunnell et al, 2020), it is likely that the pandemic will nonetheless have important influences on these behaviours. Broadly speaking these might be considered in terms of early influences, especially relating to the threat of the disease and to the public health, political and economic measures necessary to contain it and limit its effects, and then later influences, such as the impact of the pandemic on the economy, employment, finances and population mental health.

One of the most important public health measures in response to the pandemic was the introduction of lockdown restrictions, whereby individuals were asked to remain in their homes to reduce spread of COVID-19 infections. This was introduced in the UK from March 23^rd^ 2020 and continued for nearly two months, until there was some loosening of the restrictions in England after May 11^th^ 2020. A deterioration in mental health in the general population during this period has been reported in the UK (Pierce et al, 2020). Others have noted the potential mental health and psychological consequences of such public health measures (Brooks et al, 2020), including possible impacts on suicide and self-harm (Holmes et al, 2020; Niederkrotenhaler et al, 2020). Increased suicidal ideation has been reported from the USA during lockdown restrictions (Killgore et al, 2020)

Studying the factors influencing suicidal behaviour in relation to the pandemic is important as awareness of these can inform policies for prevention and intervention. This information can also be used to assist healthcare leaders responsible for service configuration and delivery and to help clinicians to be aware of key areas they should address during assessment of individuals in terms of risk or following an act of self-harm. We have investigated the factors associated with self-harm (non-fatal self-poisoning and self-injury) in adults presenting to hospital emergency departments during the first two months following the introduction of full lockdown in the UK, using data purposefully collected in two centres with well-established self-harm monitoring systems.

## METHOD

### Data collection

The study was conducted in the general hospitals in Oxford and Derby, two centres in the Multicentre Study of Self-harm in England (Geulyov et al, 2016). Information on self-harm presentations is routinely collected through long-established self-harm monitoring systems. Self-harm is defined as intentional non-fatal acts of self-poisoning or self-injury, irrespective of the type of motivation, including degree of suicidal intent (Hawton et al, 2003), the definition adopted by the National Institute for Clinical Excellence (2011). For patients who receive a psychosocial assessment conducted by mental health clinicians in the Emergency Department Psychiatric Service (Oxford) or Liaison Psychiatry Service (Derby) information is collected by the assessing clinician on data collection forms (Oxford) or electronic case records (Derby).

For the purposes of the present study, clinicians conducting psychosocial assessments were asked to complete a simple record following each assessment to indicate whether their impression was that the patient’s self-harm was influenced by any factors related to COVID-19, answering YES, UNSURE or NO. If they answered YES or UNSURE they were asked to describe the relevant factors in free text (they could indicate multiple factors if appropriate). In addition to the above information, the following items were extracted from the usual data collection form in Oxford and the electronic patient record in Derby for all patients receiving a psychosocial assessment during the study period: gender, age, date of presentation, present household composition (Oxford only), method of self-harm and, where available, score on the Suicide Intent Scale (Beck et al 1974) grouped into four categories: low (0-6), moderate (7-12), high (13-20) and very high (21 – 30).

### Classification of COVID-related factors influencing self-harm

The clinicians’ descriptions of COVID-19 related factors thought to have influenced self-harm recorded for the first 30 individuals for whom this item was recorded as positive (YES/UNSURE rating) in Oxford were initially scrutinised by KH and KL, who each independently formulated a classification of the factors. They then compared and discussed their classifications to reach a consensus on the classification system to be used in the study (see Table 2). This system was then used by the same team members in Oxford to categorise all the Oxford clinicians’ statements for patients whose self-harm was recorded to be related to COVID-19, and similarly by two team members in Derby (JN and KW) for the patients there. Any disagreements in classification were resolved through discussion between the team members at each site. Where necessary there was consultation between the Oxford and Derby teams to sort out uncertainties and ensure consistency of approach.

**Table 1.**
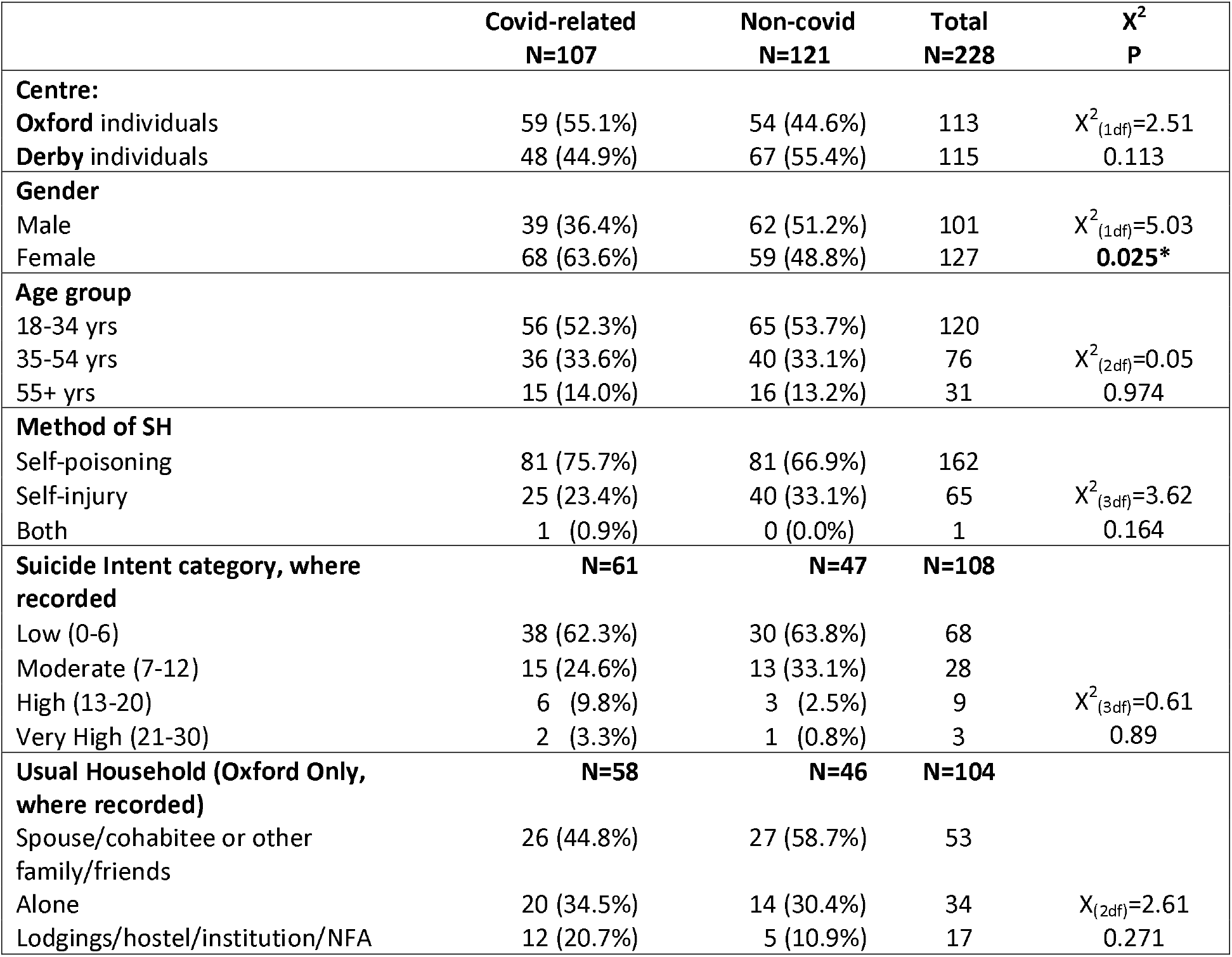
Comparison of patients in which COVID-19-related factors were and were not identified.

|  | **Covid-related**<br>**N=107** | **Non-covid**<br>**N=121** | **Total**<br>**N=228** | **$\chi^2$**<br>**P** |
| --- | --- | --- | --- | --- |
| **Centre:** |  |  |  |  |
| **Oxford** individuals | 59 (55.1%) | 54 (44.6%) | 113 | $\chi^2_{(1df)}=2.51$<br>0.113 |
| **Derby** individuals | 48 (44.9%) | 67 (55.4%) | 115 |  |
| **Gender** |  |  |  |  |
| Male | 39 (36.4%) | 62 (51.2%) | 101 | $\chi^2_{(1df)}=5.03$<br>**0.025\*** |
| Female | 68 (63.6%) | 59 (48.8%) | 127 |  |
| **Age group** |  |  |  |  |
| 18-34 yrs | 56 (52.3%) | 65 (53.7%) | 120 | $\chi^2_{(2df)}=0.05$<br>0.974 |
| 35-54 yrs | 36 (33.6%) | 40 (33.1%) | 76 |  |
| 55+ yrs | 15 (14.0%) | 16 (13.2%) | 31 |  |
| **Method of SH** |  |  |  |  |
| Self-poisoning | 81 (75.7%) | 81 (66.9%) | 162 | $\chi^2_{(3df)}=3.62$<br>0.164 |
| Self-injury | 25 (23.4%) | 40 (33.1%) | 65 |  |
| Both | 1 (0.9%) | 0 (0.0%) | 1 |  |
| **Suicide Intent category, where recorded** | **N=61** | **N=47** | **N=108** |  |
| Low (0-6) | 38 (62.3%) | 30 (63.8%) | 68 | $\chi^2_{(3df)}=0.61$<br>0.89 |
| Moderate (7-12) | 15 (24.6%) | 13 (33.1%) | 28 |  |
| High (13-20) | 6 (9.8%) | 3 (2.5%) | 9 |  |
| Very High (21-30) | 2 (3.3%) | 1 (0.8%) | 3 |  |
| **Usual Household (Oxford Only, where recorded)** | **N=58** | **N=46** | **N=104** |  |
| Spouse/cohabitee or other family/friends | 26 (44.8%) | 27 (58.7%) | 53 | $\chi^2_{(2df)}=2.61$<br>0.271 |
| Alone | 20 (34.5%) | 14 (30.4%) | 34 |  |
| Lodgings/hostel/institution/NFA | 12 (20.7%) | 5 (10.9%) | 17 |  |

**Table 2.** COVID-19-related factors identified as influencing self-harm, by gender.

| **Factors influencing Self-harm** | **Males**<br>**(N=39)** | **Females**<br>**(N=68)** | **Total**<br>**(N=107)** |
| --- | --- | --- | --- |
| **Overall mental health problems** | **11** | **22** | **33** |
| Mental health/worsening of mental health | 5 | 15 | 20 |
| Loss/reduction of supports for mental health problems | 7 | 10 | 17 |
| **Isolation /Loneliness** | **14** | **17** | **31** |
| **Lack/reduced contact** | **9** | **14** | **23** |
| Lack/ reduced contact with family | 5 | 10 | 15 |
| Reduced contact with social network | 4 | 6 | 10 |
| **Disruption to normal routine** | **6** | **14** | **20** |
| **Entrapment** | **5** | **13** | **18** |
| **Interpersonal conflict** | **3** | **9** | **12** |
| **Employment** (including loss/furloughed) | **9** | **3** | **12** |
| **Fear of COVID infection** | **3** | **7** | **10** |
| Self becoming infected | 2 | 3 | 5 |
| Self infecting others | 0 | 2 | 2 |
| Others becoming infected | 2 | 3 | 5 |
| **Accommodation/housing** | **3** | **4** | **7** |
| **Education/ training** | **1** | **6** | **7** |
| **Financial** | **5** | **1** | **6** |
| **General concerns about impact of Covid** | **0** | **5** | **5** |
| **Substance misuse** | **2** | **2** | **4** |
| Alcohol | 2 | 2 | 4 |
| Drugs | 1 | 0 | 1 |
| **Domestic abuse (actual/threatened)** | **0** | **3** | **3** |
| **Bereavement due to Covid** | **0** | **1** | **1** |
| **Other** | **2** | **2** | **4** |

### Patient Samples

The study was of patients aged age 18 years and over. In Oxford the included patients were those presenting during the eight weeks from 23^rd^ March 2020 (i.e. the first day of full lockdown in the UK) to May 17^th^ 2020. In Derby the data were for patients presenting from 2nd April to May 17^th^ 2020.

Because the study was focused on identifying COVID-19-related factors contributing to self-harm, if an individual presented with a repeat episode, the first not having been identified as related to COVID but a subsequent one having been, the latter episode was used as the index presentation for the purposes of the study. However, only one episode per person was included in the analysis.

### Data analysis

The patient characteristics and methods of self-harm were compared between those patients in which COVID-19 related factors were thought by the assessing clinician to have influenced self-harm and those in which this was not thought to be the case using the chi-squared statistic.

The data on the factors in the classification of the clinicians’ responses are presented as numbers of patients recorded as positive for each factor. Information recorded for each of the factors in the classification were explored qualitatively by KH and KL using Excel to assist identification of sub-groups within these factors.

## RESULTS

During the study period following lockdown in the UK, 228 individuals (101 males and 127 females) aged 18 years and over presented to either of the two study hospitals and received a psychosocial assessment from a mental health practitioner (113 in Oxford between March 23^rd^ and May 17^th^ 2020; 115 in Derby between April 2nd and May 17^th^ 2020). Of these, 107 (46.9%) had self-harm episodes which were considered by the clinicians as having been related to the COVID-19 pandemic in some way. This included five cases in which the clinicians had indicated ‘unsure’ regarding the contribution of the health crisis to the self-harm, but where on review of the indicated factors the research team decided that there appeared to be an association.

### Comparison of the patients with identified COVID-19-related factors contributing to self-harm and the remainder of the patients

The patients identified by the clinicians as having COVID-19-related factors influencing their self-harm are compared to the remainder of the patients in Table 1. A greater proportion of female than male patients were identified as having such factors involved in their self-harm (68/127, 53.5% v 39/101, 38.6%; χ^2^ = 5.03, p=0.025). There were no differences between the two groups of patients in terms of age groups, methods of self-harm or living circumstances (Oxford data only), and, where recorded (N=108), in the level of suicide intent.

### COVID-19-related factors contributing to self-harm

Details of the numbers of patients in which the various COVID-19-related factors were recorded as influencing their self-harm are shown in Table 2. The most frequently recorded factors were problems related to mental health, isolation/loneliness, lack of or reduced contact with other individuals, disruption to normal routine, and entrapment. It should be noted that in the majority of cases (64/107; 59.8%) more than one factor attributed to the COVID-19 pandemic was recorded, these factors often being inter-related (see below).

We now consider in more detail and in order of frequency the specific influences on self-harm identified within each of the categories of factors.

### Mental health: a) problems

The most frequent problems in this category were anxiety and depression (low mood), which in some cases were new problems and in others exacerbation of already existing conditions. In many cases they were clearly secondary to other issues, especially fear of infection or of infecting family members (usually in the absence of current physical illness). There were also single cases of onset of severe self-harm in the context of newly diagnosed psychosis in which fear of infection was a major factor, and worsening of problems related to emotionally unstable personality disorder.

### Mental health: b) loss or reduction of supports

A prominent issue in this category was reduction or cessation of support from services; for example, where the clinicians conducting the psychosocial assessments noted that because clinical support for patients had stopped due to COVID-19 their mental health had deteriorated. This included cessation of AA meetings, which resulted in one patient returning to heavy drinking

A particularly common theme was the impact of face-to-face contact with services having ceased, even when support was available by phone or other means of communication. In one case it was recorded that a likely motive for the self-harm and subsequent ED presentation was a need for face-to-face contact with a clinician. In two cases the individuals’ self-harm was related to lockdown preventing them obtaining medication prescribed for psychiatric disorders.

### Isolation/loneliness

A sense of isolation and/or loneliness resulting from lockdown and lack of contact with others was a frequent factor contributing to self-harm. In several cases it was recorded as the sole COVID-19 factor influencing self-harm. In Oxford, where data were available on living situation, 20 individuals were recorded as living alone, in nine (45.0%) of which loneliness or social isolation was identified as a factor relating to their self-harm.

### Lack of contact or reduced contact a) with family

Several patients were recorded as being affected by being unable to have face-to-face contact with their families, in some cases because they were geographically separated, even in different countries, and travel restrictions prevented access. For some individuals the lack of contact with their families was due to self-isolation either of themselves or of a family member because of health issues increasing dangers associated with infection. Loss of support from family members was a consequent contributory factor in some individuals’ self-harm. Some parents who self-harmed had particularly missed contact with their children. There were also examples of individuals being unable to have contact with a partner.

### Lack of contact or reduced contact b) with social network

The impact of lockdown on relationships with friends and social networks was an important factor in the self-harm of several individuals. This was often because of loss of support usually provided by friends.

### Disruption to normal routine

The psychological effects of having their normal routine disrupted was a factor contributing to self-harm in several patients. This included lacking a routine or structure, having nothing to do and specific disruption of plans (e.g. travel abroad, house sale).

### Entrapment

This was a relatively common contributory factor, with some patients finding their feeling trapped at home intolerable. In some this was simply identified as being unable to cope with lockdown.

### Interpersonal conflict

This was identified as a factor influencing self-harm in several individuals. It included lockdown having contributed to worsening of a previously difficult relationship with a partner or a relationship coming under strain, conflict or tension between family members, and breakup of relationships with partners.

### Employment

COVID-19-related employment issues were identified by the clinicians as contributing to self-harm in some patients. These included individuals who were unemployed but unable to seek work because of lockdown, others who had become unemployed because of the pandemic, individuals who had been unable to continue their usual work due to vulnerability resulting from physical health problems or underlying health issues necessitating shielding, and some whose self-harm was related to being furloughed.

### Fear of infection

The most common infection fear identified as contributing to self-harm was about individuals themselves becoming infected with COVID-19. In a couple of cases the fear was extreme, one individual believing that if they got infected they would die and therefore might as well kill themself first. Another patient greatly changed their eating habits as they thought this could provide protection against infection. Two patients feared that if they became infected with COVID-19 they would transmit this to vulnerable relatives.

Some patients had general concerns about family members becoming infected, either due to their working on the acute healthcare front line or because of other specific vulnerability.

### Accommodation/housing

There were several individuals where problems related to accommodation or housing resulting from restrictions due to the pandemic were thought to have influenced their self-harm. This was for a variety of reasons, including being stuck in student accommodation or unsuitable housing, and being unable to move into more suitable accommodation.

### Education/training

This was recorded as a factor in a few cases, in all of which other factors were also identified. For some, it was the inability to continue educational studies and in one the stress of providing home schooling.

### Financial Problems

These were either worries about finances or an actual lack of funds, usually secondary to lockdown, mostly because of loss of a job or inability to get work.

### General concerns about the COVID-19 pandemic

A few patients were identified as having general concerns about the impact of the pandemic, including worrying about the numbers of people who had died.

### Alcohol/drug misuse

In the few cases in which alcohol misuse was considered a pandemic-related factor during lockdown this involved increased drinking, including cessation of a long period of abstinence in one individual with alcohol use disorder, and another who contravened lockdown restrictions to access both alcohol and drugs.

### Domestic Abuse

While not a common contributory factor, it was recorded for some female patients who felt threatened by previously violent male partners.

### Bereavement due to COVID-19 infection

In a single case the key factor identified as associated with self-harm was death of a partner from COVID-19 infection.

### Other problems

There was a small number of patients where the factors identified did not fit in one of the above categories. These included sleep disturbance, boredom and struggling to cope as a carer for family members,

### Inter-relationship between factors

For many patients there were multiple inter-related factors resulting from the COVID- 19 pandemic that had contributed to their self-harm. The following are examples (details modified to preserve anonymity):

> A patient with mental health problems was anxious about leaving their house because of risk of infection. Their contact with their parents had been much reduced. A care assistant was no longer visiting and face-to-contacts with a CPN and a therapist was now only available over the phone.
>
> A patient whose child and another relative had been infected with COVID had subsequently become increasingly depressed and anxious. Their partner worked in a care setting where personal protective equipment had been limited, which further increased their anxiety, and added to their sense of isolation and low mood.
>
> A patient with mental health problems living with a volatile partner found their constantly being together at home and consequent arguments increasingly stressful. They felt trapped, a feeling that was exacerbated by a delay in provision of previously planned treatment for their psychiatric problems.

## DISCUSSION

We have used data collected through two well-established hospital-based self-harm monitoring systems and information recorded by clinicians based on their psycho- social assessments conducted with patients to investigate the extent to which the COVID-19 pandemic may have influenced self-harm, and the factors that contributed to self-harm during the two months following introduction of lockdown restrictions.

The clinicians’ assessments indicated that COVID-19-related factors influenced self-harm for nearly half of the patients. This was the case for 53.5% of the females compared with 38.6% of the males, suggesting that lockdown may have been more stressful for females. However, there were no differences between those whose self-harm were assessed as being related to COVID-19 compared with those for whom it was not in terms of age, methods of self-harm or suicide intent. This suggests that the severity of self-harm did not differ between the two groups of patients, either physically or psychologically.

The most frequent COVID-19-related factors recorded by clinicians concerned mental health. These included onset or exacerbation of mental health problems and reduced availability of services. The latter included inability to have face-to-face contact with clinicians. This suggests that there must be care in remodeling psychiatric services because of the pandemic (Moreno et al, 2020), including ensuring that remote working is not regarded as the norm and that the needs and risks of individual patients are taken into account.

The next most frequent COVID-19-related factors concerned those likely to be directly related to lockdown restrictions, namely isolation and loneliness, reduced contact with families and friends, disruption of normal routine and a sense of entrapment. These findings are largely in keeping with predicted psychological impacts of lockdown (Holmes et al, 2020). They highlight the need for relatives, friends and neighbours to be encouraged to reach out to others, especially those living alone, to help provide a sense of socialisation and support, including through telephone and internet communication where face-to-face contact is not possible.

While some expected consequences of the COVID-19 pandemic such as employment and financial problems (Gunnell et al 2020; Niederkrotenhaler, 2020) were not frequently identified in this series of patients, this is likely to be due to the focus having been on the early stages of the pandemic, together with the impact of UK government initiatives to protect jobs through furloughing and other financial supports. Economic and financial factors are likely to become more influential as the longer term impacts of the pandemic develop.

Multiple COVID-19-related factors were often identified as influencing self-harm in individual cases, with specific factors such as isolation, entrapment and interpersonal difficulties seeming to initiate a sequence of other influences related to the pandemic. Reduced access the mental health services and other means of support were often part of the matrix of factors.

The findings of this study have implications for clinical practice and the organisation of services during the pandemic and its aftermath. They highlight factors that clinicians assessing patients who have self-harmed can be sensitive to during their assessments. The categorisation of factors we have developed might be used as an aide-memoire while conducting assessments and safety planning (a blank version is included as an online supplement to assist with this). We have already noted that a cautious approach should be taken to deciding who, and who may not, be suitable for remote clinical care for psychiatric problems, although of course physical safety of both patients and staff must be a priority consideration. Every effort should be made to sustain mental health services in a way that is responsive to the needs of individuals and the community, even during very difficult circumstances (Moreno et al, 2020). Our findings are not just relevant historically, but also to potential developments of this and future pandemics, including necessary reintroduction of stringent public health measures to address recurrence of disease spread.

The study methodology and findings also have relevance for future research. Our classification, which was based on clinicians’ impressions, might be developed into an interview schedule. It will be important to see whether influences of the pandemic change with time, especially as longer-term consequences develop. Research regarding both clinician and service user perspectives of provision of clinical care under the restrictions consequent on the pandemic and resulting public health measures is clearly required in order to optimise clinical interventions.

## Strengths and limitations

One strength of the study is that it was conducted in two centres with very different catchment area population characteristics in terms of levels of socio-economic deprivation (Geulyov et al 2016). Also, the clinicians’ responses regarding the COVID-19-related factors influencing self-harm were spontaneous and not influenced by a structured questionnaire, although this may also have meant that some relevant factors were not identified.

Limitations include the fact that the patients in the study were of necessity only those who received a clinical assessment. While the rates of assessment are very high in both centres compared with those found nationally (Cooper et al, 2013), it is recognised that patients who do not receive an assessment may differ from those who do with regard to gender, method of self-harm and other characteristics such as alcohol use (Hickey et al 2001; Bennewith et al 2005; Kapur et al 2008), which could also apply to whether the pandemic influenced their self-harm. However, we did not find any difference in the methods of self-harm used by the assessed patients with and without COVID-19 factors influencing their self-harm. Finally, scores on the Suicide Intent Scale were not available for a substantial proportion of patients.

## Conclusions

More frequent factors related to the COVID-19 pandemic which were assessed by clinicians as having influenced self-harm included mental health problems and restricted mental health service provision, isolation and loneliness, reduced contact with friends or family members, disruption of normal routine and entrapment. The results have implications for mental health service provision and social responses to the pandemic and consequent public health measures. The findings are relevant to future responses to the pandemic, especially if and where lockdown is reintroduced. The classification developed during the study can help clinicians who are assessing people who have self-harmed and conducting safety planning. It could also be developed for future research investigations.

## Data Availability

The data for this study are not available for sharing because they are collected without patient consent and the approval agency for this does not allowus to share the data.

## Contributors

KH and KL were responsible for study conception and design, and interpretation of the results. KH, KR and DC were responsible for data analysis. LB, FB, JN, KW and SK acquired the data. KH and KL drafted the report, which all authors critically revised for intellectual content. All authors approved the final report and are accountable for all aspects of this work. KH supervised the study and is the guarantor.

## Acknowledgements

The study was funded by the Department of Health and Social Care. KH is supported by Oxford Health NHS Foundation Trust.

We thank the clinicians in the Emergency Department Psychiatric Service in Oxford and the Liaison Psychiatry Service in Derby and the research staff in both centres for their support with the data collection. The authors from Derby would like to thank Jessica Pearson and Phyllis Leung (Research Assistants).

## Online Appendix

### COVID-19-related factors that may influence self-harm Aide-memoire to assist clinicians assessing patients

Aide-memoire to assist clinicians assessing patients
|  |
| --- |
| **Factors influencing self-harm** |
| **Overall mental health problems**<br>Mental health/worsening of mental health<br>Loss/reduction of supports for mental health problems |
| **Isolation /Loneliness** |
| **Lack/reduced contact**<br>Lack/ reduced contact with family<br>Reduced contact with social network |
| **Disruption to normal routine** |
| **Entrapment** |
| **Interpersonal conflict** |
| **Employment** (including loss/furloughed) |
| **Fear of COVID infection**<br>Self becoming infected<br>Self infecting others<br>Others becoming infected |
| **Accommodation/housing** |
| **Education/ training** |
| **Financial** |
| **General concerns about impact of Covid** |
| **Substance misuse**<br>Alcohol<br>Drugs |
| **Domestic abuse (actual/threatened)** |
| **Bereavement due to Covid** |
| **Other** |

